# Computational Strategies in Nutrigenetics: Constructing a Reference Dataset of Nutrition-Associated Genetic Polymorphisms

**DOI:** 10.1101/2023.08.04.23293659

**Authors:** Giovanni Maria De Filippis, Maria Monticelli, Alessandra Pollice, Tiziana Angrisano, Bruno Hay Mele, Viola Calabrò

**Author notes:** **Correspondence to:** Bruno Hay Mele, Biology Dept., University of Naples Federico II, Via Cinthia, 26 Naples, Italy.

## Abstract

**Objective:** This study aims to create a comprehensive dataset of human genetic polymorphisms associated with nutrition by integrating data from multiple sources, including the LitVar database, PubMed, and the GWAS catalog. This consolidated resource is intended to facilitate research in nutrigenetics by providing a reliable foundation to explore genetic polymorphisms linked to nutrition-related traits.

**Methods:** We developed a data integration pipeline to assemble and analyze the dataset. The pipeline performs data retrieval from LitVar and PubMed, data merging to build a unified dataset, definition of comprehensive MeSH queries in order to retrieve relevant genetic associations, and cross-referencing the results with the GWAS data.

**Results:** The resulting dataset aggregates extensive information on genetic polymorphisms and nutrition-related traits. Through MeSH query, we identified key genes and SNPs associated with nutrition-related traits. Cross-referencing with GWAS data provided insights on potential effects or risk alleles associated with this genetic polymorphisms. The co-occurrence analysis revealed meaningful gene-diet interactions, advancing personalized nutrition and nutrigenomics research.

**Conclusion:** The dataset presented in this study consolidates and organizes information on genetic polymorphisms associated with nutrition, facilitating detailed exploration of gene-diet interactions. This resource advances personalized nutrition interventions and nutrigenomics research. The dataset is publicly accessible at https://zenodo.org/records/14052302, its adaptable structure ensures applicability in a broad range of genetic investigations.

## 1 Introduction

Nutrition is critical to health and disease [1]. Emerging evidence suggests that genetic polymorphisms significantly impact an individual’s response to different nutrients and dietary patterns by affecting nutrient bioavailability and metabolism [2]. Moreover, it has been demonstrated that common gene variations are linked to complex chronic health issues significantly affected by nutritional factors [3].

Advancements in genomics technologies and the subsequent availability of large-scale genetic data have fueled interest in the identification and categorizing of genetic polymorphisms associated with nutritional traits [4]. Thus, the field of nutritional genetics (nutrigenetics) was born to comprehend how genetic variations influence an individual’s nutritional requirements, metabolism, and health outcomes [5]. By considering an individual’s genetic profile, healthcare professionals and nutritionists can provide tailored dietary advice and interventions that optimize nutrient bio-availability and promote better health outcomes in that individual [6]. Nutrigenetic associations imply that specific genetic polymorphisms can induce susceptibility to chronic diseases. The response to specific nutrients or dietary patterns may be crucial in determining health outcomes [7]. Recent literature contains extensive data on nutrition-associated genetic polymorphisms [2, 7]. However, these data are often scattered, diverse in format, and lack a standardized curation process. Such complications hinder data integration, limit information extraction and synthesis, and pose a barrier to data utilization in decision support systems [8].

Integrating available data and overcoming the limits of self-reported methods in research is crucial for accurate *omics data integration, nutrigenetics, and nutrigenomics research, especially in clinical settings [8]. Therefore, there is a need to develop comprehensive and structured resources that integrate nutrition-associated genetic polymorphism data, along with *omics data, to advance personalized nutrition interventions and clinical decision-making. Today, technologies are available to overcome these limitations: the use of ontologies for information retrieval (IR) is a well-known technique in the literature for semantic search [9], while Named Entity Recognition (NER) techniques are increasingly important in biomedical literature mining [10] to obtain key information on genomic variants for personalised medicine.

Here, we built a structured dataset of human genetic polymorphisms associated with nutrition by mining the LitVar database [11], which contains curated information on genetic variations and their functional effects; the PubMed-Medline database, which provides structured MeSH ontology annotations; and the GWAS catalog dataset, which reports human variant-traits associations. Our dataset includes data from PubMed studies associated with nutrition-related genetic polymorphism. This data where then queried employing MeSH ontology for retrieval of nutrition-related genetic data. Specific sets of MeSH terms related to nutrition physiology, nutrition-related diseases, prevention through diet, and eating behavior were used to retrieve subsets of genes and their single-nucleotide polymorphisms (SNPs) potentially associated with nutrition-related traits. Cross-referencing with the GWAS catalog dataset [12] provided information about effect/risk alleles associated with the collected studies. The resulting dataset was validated to ensure data quality, consistency, and relevance to nutrition and nutrigenomics research, thus providing a valuable resource to investigate the intricate interplay between genetics and nutrition.

## 2 Methods

We developed an integrated dataset by cross-referencing genomic data with scientific literature using shared PubMed IDs to link the LitVar and PubMed databases. This linkage allowed us to enrich LitVar’s association data by incorporating Medical Subject Headings (MeSH).

The *MeSH Ontology* is a structured and controlled vocabulary that supports the annotation and indexing of biomedical literature and datasets. It provides standardized descriptors that facilitate the organization and retrieval of scientific information within the domains of biomedicine and healthcare informatics. The structured nature of MeSH terms enhances their applicability and robustness for comprehensive information retrieval and analysis of scientific indicators. In a database context, MeSH terms function as metadata references, enabling precise literature categorization and search [13].

The LitVar database is an extensive and publicly accessible repository that aggregates information on genetic variants and links them with corresponding scientific literature. Its purpose is to address the challenge of connecting genomic data with the relevant literature by synthesizing information on genetic variants from diverse sources, employing NER techniques [11]. PubMed operates as the primary digital library for biomedical literature, offering an invaluable resource for scientific inquiry. Data extraction from PubMed is essential for various research activities, including systematic reviews, data mining, and knowledge discovery [14].

To construct our dataset, we implemented a Python-based data processing pipeline designed to integrate heterogeneous data sources by utilizing the MeSH ontology as a central schema for data harmonization. This pipeline allows extraction, integration, and querying of genetic polymorphism information pertinent to specific research domains, such as nutrigenetics. The output dataset, referred to as GRPM dataset, encapsulates primary identifiers, including Genes, Reference SNP IDs (RsIDs), PubMed IDs (PMIDs), and MeSH terms. Recognizing the pivotal role of genetic influences in nutrition-related traits, the GRPM dataset facilitates comprehensive exploration and analysis, thereby supporting researchers and nutritionists in advancing personalized nutrition studies.

The data retrieval and integration pipeline is implemented in a Jupyter Notebook environment [15] and comprises five distinct modules, each tailored to perform specific functions essential for the assembly and utilization of a comprehensive genetic polymorphism dataset (as illustrated in Figure 1). These modules are outlined as follows:

**Figure 1:**
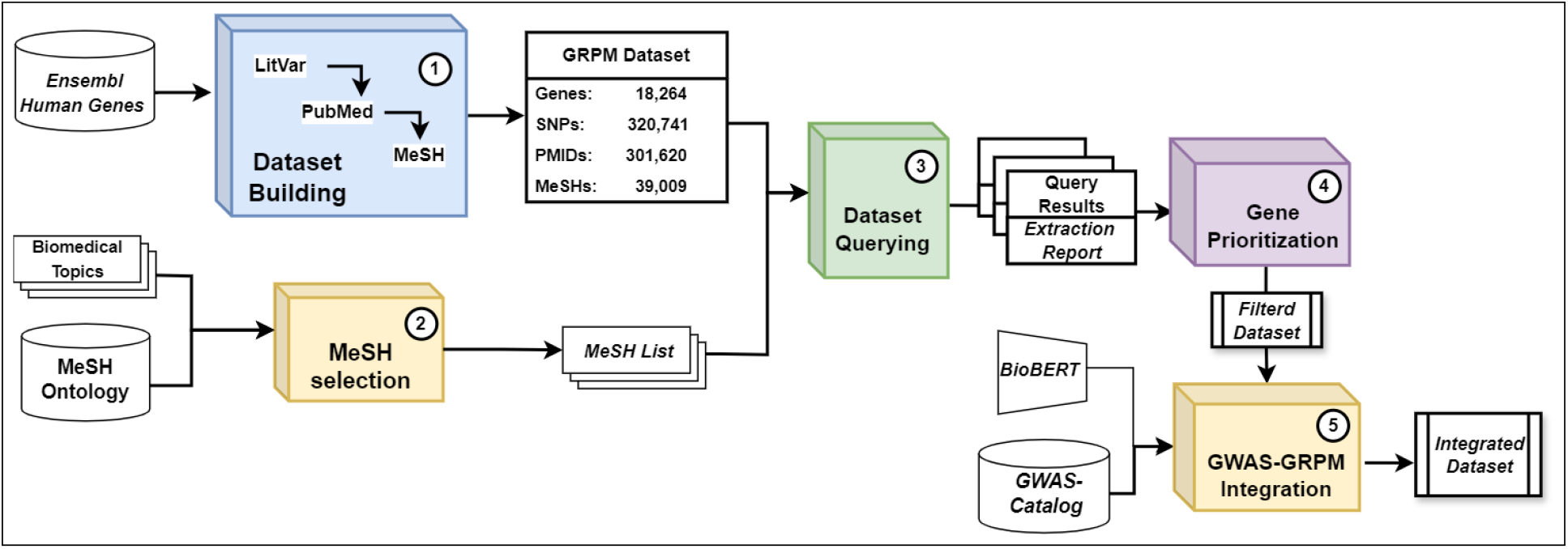
Graphical overview of our workflow showcasing the input data and interactions between the five modules.

1. *GRPM Dataset Building*: This module orchestrates the extraction, integration, and consolidation of data from source databases, including LitVar and PubMed. It ensures the comprehensive collection of genetic polymorphisms associated with topic-related traits (GRPM dataset).
2. *MeSH Term Selection*: This module facilitates the definition of MeSH ontology term sets. These are employed to query the GRPM dataset.
3. *GRPM Dataset Querying*: This module allows users to query the dateset using predefined MeSH terms. Users can thus refine their searches to focus on specific areas of interest, ensuring targeted data retrieval pertinent to their research needs within the GRPM dataset.
4. *Gene Prioritization*: This module assigns a relative importance score to each gene based on the frequency and proportion of associated findings. This metric enables the prioritization of genes pertinent to specific research topics, aiding further investigation.
5. *GWAS Data Integration*: The integration of the GWAS catalog provides additional insights into effect and risk alleles associated with the identified genetic polymorphisms. This process enriches our nutrigenetic resource with supplemental data for further analysis.

All implementation details, along with usage instructions, are available in the open-access repository on GitHub^1^.

### 2.1 GRPM Dataset Building

The first module uses the LitVar Application Programming Interface (API)^2^ to retrieve all polymorphisms for each human gene within the LitVar database alongside all associated PMIDs. These PMIDs were subsequently employed as queries on PubMed to obtain the bibliographical data. We empoloyed an NBIB parser^3^, to extract and structure this data in a machine readable format. The collected data were ultimately consolidated into a single CSV file (“GRPM dataset”), serving as the primary source against which MeSH term queries can be employed to retrieve genes and polymorphisms associated with specific contexts.

This work is based on the first version of LitVar, which is no longer available online and has been entirely replaced by LitVar 2.0 [10]. This version was chosen based on several reasons. Firstly, the first version of LitVar possesses a higher level of reliability, a product of extensive examination and rectification of any discrepancies over its period of usage. Besides, the relatively simpler structure of the data in this version eschews unnecessary complexity posed by more recent data structures, thereby making data extraction and manipulation operations more straightforward. The decision to use LitVar first version was the result of a thorough cost-benefit analysis, weighing the potential superior data precision provided by LitVar 2.0, which also comes with substantially larger datasets that could introduce additional noise, against the reliability and simplicity of the first version. The dataset produced here provides a faithful and historical archive of the first version of LitVar by collating the bibliographic references along with the genes and polymorphisms associated with them.

### 2.2 Query definition

The retrieval system to get subsets of genes and polymorphisms from GRPM dataset employes a user-defined list of MeSH terms as a hook. Careful selection of the MeSHs is crucial at this stage: the list must represent the chosen search field out of the total complex of terms in GRPM dataset. The total set of MeSH describing the GRPM dataset comprises 21,705 terms related to LitVar publications retrieved from the complete MeSH ontology^4^ (348,733 terms). Therefore, this subset collects ontology terms linked to papers exploring the associations between genetic variants and biomedical traits.

The second module is designed to select from the MeSH ontology a set of terms that represent a specific biomedical fields. Initially, we constructed a collection of topic-specific terms based on our domain knowledge in nutrigenetics. These topic-relevant terms were then processed to identify corresponding MeSH terms using a straightforward Natural Language Processing (NLP) approach. Subsequently, the extracted MeSH terms underwent a filtering process based on their defined “semantic types”. This step was followed by a manual screening to remove any ambiguous or potentially biased terms. This rigorous filtration ensures that only the most relevant terms are employed for dataset querying.

### 2.3 GRPM Dataset querying

In the third module, the predefined MeSH list is utilized to execute targeted queries on the GRPM Dataset, aimed at retrieving association data pertinent to genetic research (*“Survey Dataset”*).

Upon execution of the queries, an extraction report is compiled, aggregating the occurrence statistics from various data sources (NCBI, LitVar and PubMed) for each retrieved gene. These reports are subsequently subjected to both individual and comparative analyses in the fourth module.

### 2.4 Nutrigenetic Dataset Building

In order to build a nutrigenetic dataset, we defined ten major topic of interest covering nutrition physiology, nutrition-related diseases, disease prevention through diet, and eating behavior. For each of these topics, following the procedure described in the 2.2 section, we prepared MeSH queries to submit to our dataset. The topics delineated include:

1. *General Nutrition*: Encompasses a broad range of issues concerning dietary patterns, nutritional status, and overall health.
2. *Obesity, Weight Control, and Compulsive Eating*: Focuses on weight management and related disorders.
3. *Cardiovascular Health and Lipid Metabolism*: Related to the impact of diet on heart health and lipid levels.
4. *Diabetes Mellitus Type II and Metabolic Syndrome*: Covers dietary interventions and metabolic complications.
5. *Vitamin and Micronutrients Metabolism and Deficiency-Related Diseases*: Involves the metabolism and health impact of vitamins and micronutrients.
6. *Eating Behavior and Taste Sensation*: Related to food choices, taste preferences, and appetite regulation.
7. *Food Intolerances*: Addresses adverse reactions to specific foods and their genetic basis.
8. *Food Allergies*: Explores genetic aspects and dietary management strategies for allergies.
9. *Diet-induced Oxidative Stress*: Investigates how diet influences oxidative stress and health outcomes.
10. *Xenobiotics Metabolism*: Focuses on how the body processes foreign substances like drugs and toxins.

For more detailed descriptions and the MeSH count associated with each topic, please refer to Table S1 in the Supplementary Materials.

The nutrigent dataset was then constructed by merging the results of the 10 queries and filtering them using a metric based on the number and specificity of publications associated with each gene. Then, we calculated for each gene a score (**Gene Interest Index (GI)**), considering potentially “interesting” a gene if its related SNPs are associated with a substantial number of PMIDs (PubMed IDs) that include MeSH terms in the query and if the ratio between these PMIDs count and the total number of gene-associated PMIDs is sufficiently high.

To evaluate the pertinence of the retrieved gene set for the specified topic, it is essential to treat the employed MeSH set as a cohesive unit rather than analyzing the terms in isolation. This approach acknowledges the varying significance of each term within the context of the overall ontology. To determine if a gene is “interesting” based on its linked MeSH terms in LitVar-annotated studies, we propose normalizing the count of identified PMIDs by the total PMID count associated with that gene within the LitVar database. This methodology aids in reducing selection bias that may arise from genes that are extensively researched and thus associated with a larger number of MeSH terms, which might not be directly relevant to the specific query at hand.

Given the set of genes *L*(*i*) retrieved with the query (*j*), we introduce the following indices:

1. *P*_*gi*_: The total number of PMIDs associated with gene *i*;
2. *P*_*mi,j*_: The number of *i*-related PMIDs containing at least one MeSH from the query *j*;
3. *P*_*mmax*_: The highest *P*_*mi,j*_ value across all the genes in *L*;
4. *P*_*mscorei,j*_: the *P*_*mi,j*_ value normalized *P*_*mmax*_;
5. *P*_*mratioi,j*_: the ratio of *P*_*m*_ to *P*_*g*_. It measures the proportion of matching PMIDs to the total PMIDs associated with the gene.

Based on these indices, we introduce the *“Gene Interest Value”* (GV), calculated as the product of “*P*_*m*_ score” and “*P*_*m*_ ratio” and its normalized form, the **“Gene Interest Index”** (GI), which is adjusted relative to the maximum value obtained in the survey. The ratio serves as a modifier in determining the level of interest for each gene.

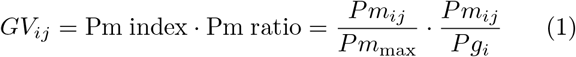

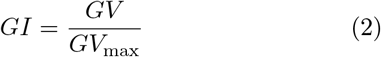

By integrating the *P*_*m*_ score and *P*_*m*_ ratio, the GI method acts as a coherent measure of gene relevance. Figure 2 visually represents an example of gene prioritization obtained through the Index using the “Obesity and Weight Control” MeSH list as a reference. Panel (a) shows the *P*_*m*_ ratio (green) and *P*_*m*_ score (yellow). It highlights the importance of considering both indexes, which produce different orders. In Panel (b), the gene relevance-based sorting achieved with the GI is presented, and it is possible to appreciate the highest prioritization performance versus the other two. The integrated assessment provided by the GI method allows for more accurate gene prioritization, leading to a deeper understanding of gene-gene interactions and potential therapeutic targets in obesity and weight control management. Another example of gene prioritization through GI is presented in Supplementary Materials (Figure S1).

**Figure 2:**
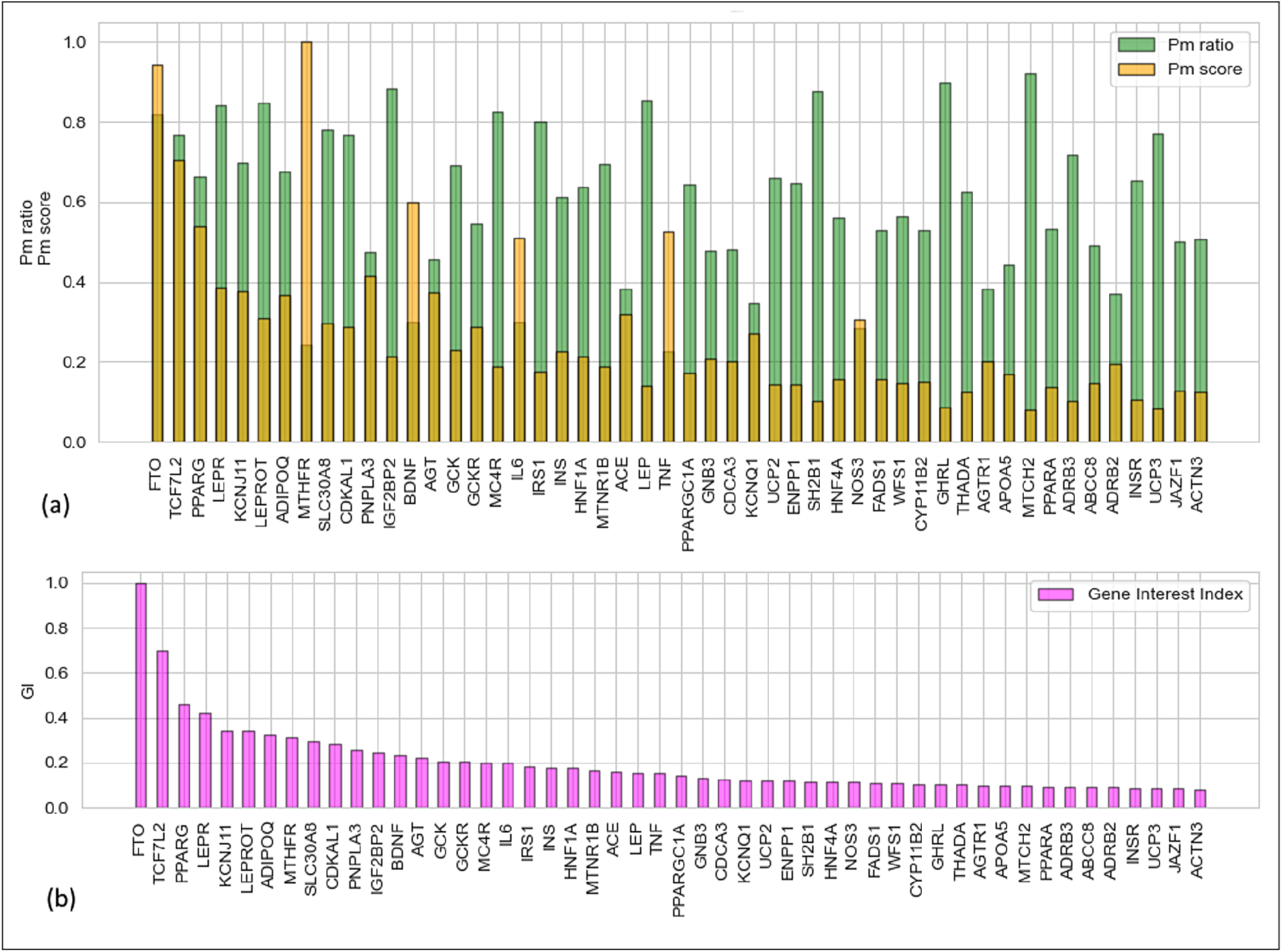
Visual representation of Gene Interest Index (GI) calculation though *P*_*m*_ score and *P*_*m*_ ratio, using as reference the results from *“Obesity and Weight Control”* MeSH query (see results section). Panel (a) shows the matching PMID ratio and overall matching PMID score. Panel (b) displays the gene relevance sort achieved with the GI.

In Section 3, we present the results obtained by applying the Gene Interest Index (GI) to ten nutritional MeSH queries results for genetic association retrieval on the GRPM dataset. We established a GI cut-off of 0.0125, which corresponds to the mean value of the 95th percentile across all ten query results. This threshold encompasses the top 5% of the retrieval results on average, thereby accommodating the long-tail distribution characteristic of the data.

Most protein-coding genes had citations with at least one of the MeSH in the query, but not all are relevant. By setting a GI threshold, we prioritized genes that fit our tailored MeSH terms, focusing on those with higher relevance in nutrigenetic dietary advice. This helped eliminate noise and focus on genes likely to offer valuable insights into gene-diet interactions and personalized nutrition.

### 2.5 GWAS data integration

The exploration of each study to discern the associated effect allele for every SNP ID presents a challenge due to its time-intensive nature. An initial assessment of the potential effect allele is instrumental for conducting preliminary investigations. To streamline this process, we integrated the Ensembl GWAS Catalog data^5^ [12] into our nutrigenetics dataset, in the fifth module in our data pipeline.

To incorporate GWAS data, we leveraged the BioBERT language model, a state-of-the-art tool specifically tailored for biomedical text mining [16]. BioBERT, an adaptation of BERT, is designed to capture and represent complex biomedical concepts within a high-dimensional vector space. This enhanced representation is instrumental in discerning semantic relationships within biomedical terminology. This approach for semantic annotation and entity linkage draws on methodologies similar to those evaluated in Tutubalina et al.’s work on information extraction using BERT models for biomedical search engines [17]. We employed BioBERT-generated embeddings for both MeSH terms and GWAS trait descriptions to assess semantic similarity. This was achieved by calculating the cosine similarity between these numerical vectors, allowing for the identification of closely related entities. Only associations that scored above a 90% similarity threshold were retained, following a thorough manual verification of their correctness. This step provided us with a reliable correlation map between MeSH concepts and GWAS traits or diseases. Subsequently, we merged the enriched GWAS Catalog data with our nutrigenetic dataset, aligning them based on SNP identifiers. The integration was further processed using the correlation map, ensuring that only highly relevant associations were included. As result, the enhanced dataset includes information on the most significant SNP-risk alleles linked to each Reference SNP ID (rsID).

Additionally, the alignment of GWAS associations acts as a quality validation mechanism for the proposed methodology based on MeSH terms, counteracting potential biases often introduced by text mining. The inclusion of a MeSH term does not inherently verify that the cited SNP is biologically associated with the queried phenotype; rather, it ensures citation within the same study, securing more accurate phenotype-genotype connections. By incorporating GWAS data, we can mitigate these biases, ensuring that the associations identified through MeSH-driven queries align with validated genetic correlations within the domain of personalized nutrition research.

## 3 Results

As result, using the outlined workflow, we have constructed a primary dataset, termed GRPM, from which we have extracted a nutrigenetics-specific subset using custom-designed MeSH-based queries. This dataset was filtered using the GI metric to select the most relevant results. Subsequently, it was augmented with GWAS data through semantic integration techniques. Consequently, the outcomes are three datasets of progressively smaller size:

1. *Primary GRPM Dataset*: Dimensions (16,610,132 rows, 6 columns)
2. *Nutrigenetic Subset*: Dimensions (1,171,249 rows, 6 columns)
3. *Nutrigenetic Dataset Augmented with GWAS*: Dimensions (179,664 rows, 15 columns)

In Tables 1 through 2 and 3, examples of entries for the three datasets along with their respective data dictionaries are provided. These datasets are stored on Zenodo (https://zenodo.org/records/14052302) and can be easily accessed using the provided Jupyter Notebooks or by installing the Python package (https://github.com/johndef64/GRPM_system). Query examples can be found in the *‘test’* directory of the package.

**Table 1:** GRPM Dataset description.

| Data Dictionary |  |  |  |
| --- | --- | --- | --- |
| Field | Description | Type | Example |
| gene | Gene symbol | string | GLIS3 |
| type | Gene type* | string | PCG |
| rsid | Reference SNP ID | string | rs779079998 |
| pmid | PubMed ID for associated literature | string | 17076841 |
| mesh | Medical Subject Headings | string | "Diabetes Mellitus, Type 2" |
| qualifier | Qualifier for MeSH term | string | genetics |
| major | Indicates if the MeSH term is a major topic | boolean | True |

| Dataset Sample entry |  |  |  |  |  |  |
| --- | --- | --- | --- | --- | --- | --- |
| gene | type | rsid | pmid | mesh | qualifier | major |
| DNMT3A | PCG | rs377577594 | 25315154 | DNA Methylation | drug effects | True |
| GLIS3 | PCG | rs7020673 | 24060607 | Diabetes Mellitus, Type 2 | genetics | True |
| CFB | PCG | rs779079998 | 25222132 | NIH 3T3 Cells |  | False |
| EYA1 | PCG | rs3779747 | 23840632 | Protein Tyrosine Phosphatases | metabolism | False |
| ABCB1 | PCG | rs2032582 | 33326171 | Ovariectomy |  | False |

\* PCG: protein coding genes; RNA: RNA genes; PSD: pseudogenes

**Listing 1.**
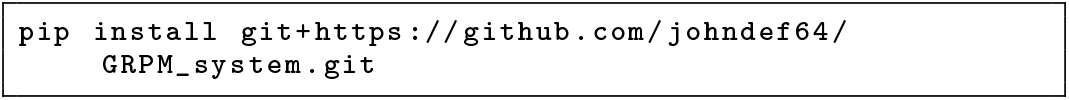
Python package installation

LitVar is a significant resource developed through extensive NER of genetic literature. However, the Lit-Var API (https://www.ncbi.nlm.nih.gov/research/litvar2/api) currently supports queries using gene symbols or variants (rsID), but does not facilitate retrieval based on phenotypic effects. Our research addresses this limitation by enhancing the LitVar dataset with MeSH ontology and GWAS phenotypes. This integration facilitates targeted retrieval of variants from indexed literature, enabling the construction of a specialized nutrigenetics dataset.

### GRPM Dataset

Figure 3 presents a compositional and comparative overview of the primary GRPM dataset. This dataset comprises approximately 77% of the PMIDs retrieved from the LitVar database (as of June 2023).

**Figure 3:**
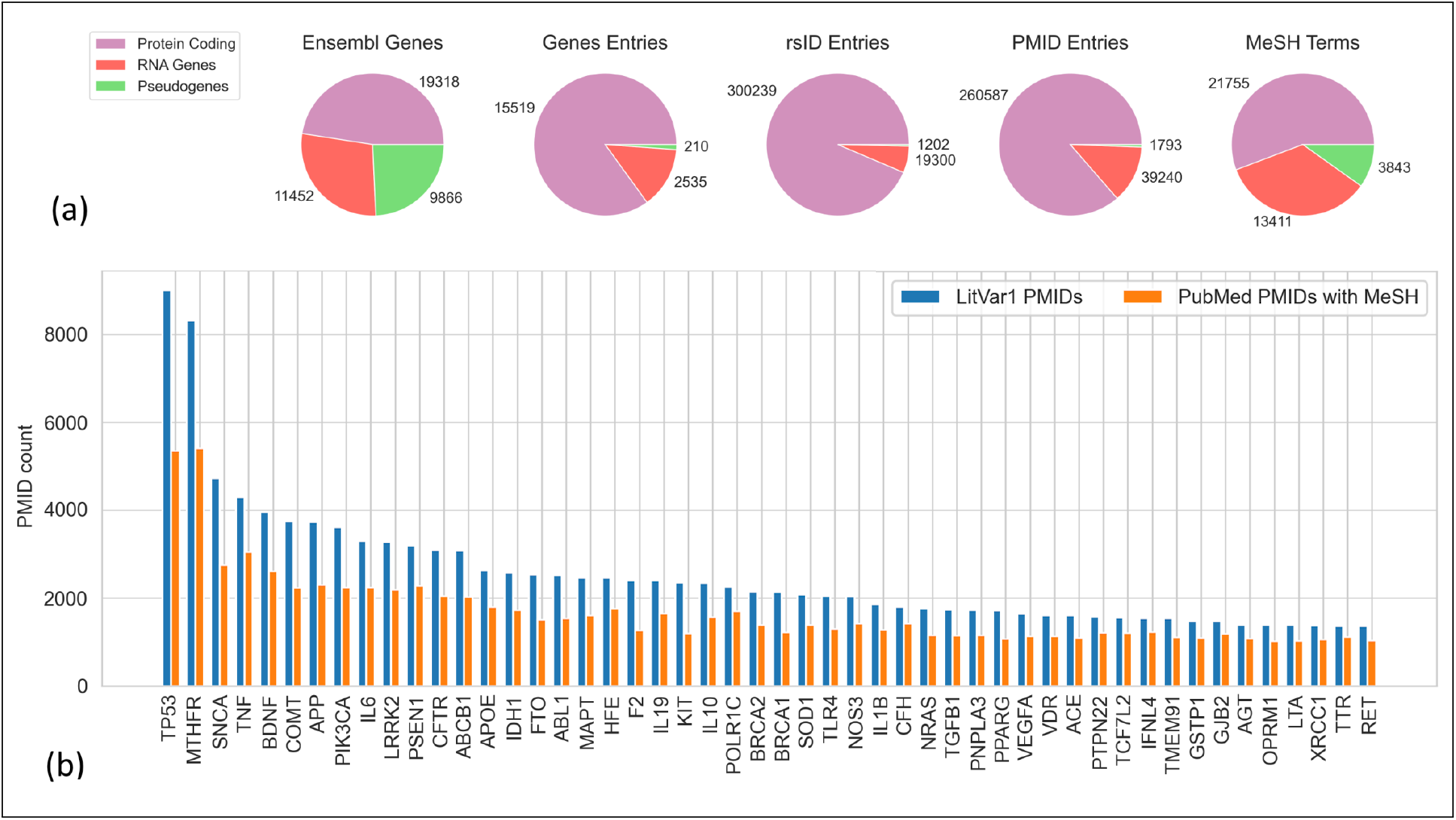
Overview of the Primary GRPM Dataset. Panel (a) shows the distribution patterns of genes, rsID, PMID, and MeSH terms for each entry type. Panel (b) identifies the 50 most frequent genes within the dataset, alongside their referenced PMID counts and correlated MeSH terms.

These PMIDs have been associated with MeSH terms sourced from PubMed, facilitating enhanced semantic annotation and interoperability.

Employing this dataset, it is crucial to consider the PMID count associated with each gene. Genes with higher research prominence are linked to a greater number of PMIDs, resulting in an increased number of MeSH annotations. Therefore, to ensure proper data normalization and analysis, it is vital to comprehensively identify and understand the most frequently represented genes within the dataset (Figure 3b).

### Nutrigenetic dataset

Our study aimed to create a *nutrigenetic dataset* using a collection of MeSH terms related to different aspects of nutrition.

Table 2 provides a statistical overview of the data sourced from nutritional MeSH-based queries executed on the GRPM dataset. Given the extensive scope of our dataset, which includes a wide range of genes and associated MeSH terms, identifying genes that are most pertinent to specific research objectives presents a notable challenge. To address this complexity, this primary results are refined using a Gene Interest (GI) metric threshold of 0.0125. This filtering criterion is crucial for isolating the most significant genetic associations, thereby enhancing their utility in nutrigenetics research. By leveraging this data-driven approach, we have effectively prioritized the extraction of the most pertinent and impactful data, facilitating the development of our final, refined dataset.

**Table 2:**
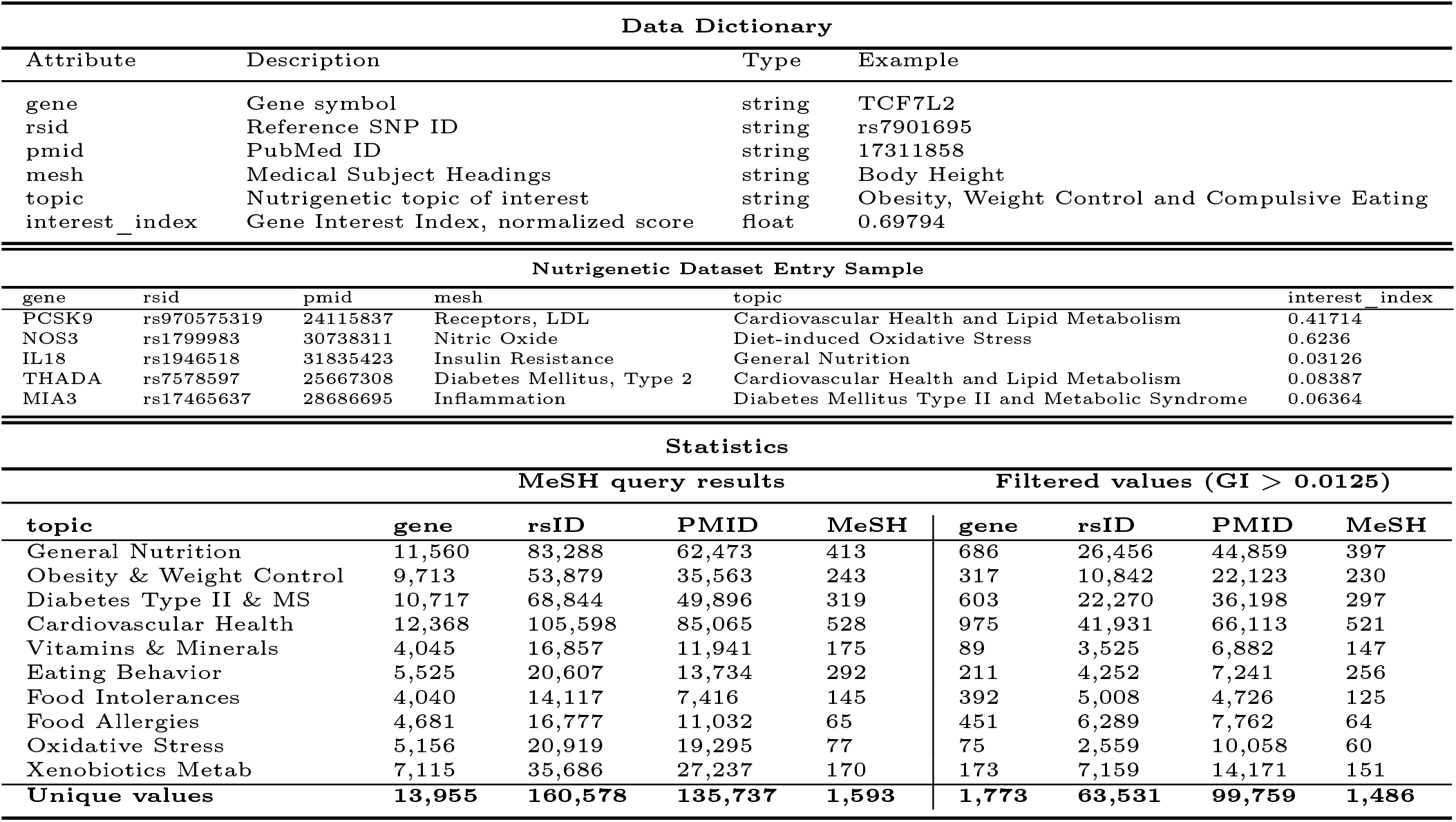
Nutrigenetic Datasest structure and statistics.

### Nutrigenetic GWAS dataset

Cross-referencing data between our refined nutrigentic dataset and GWAS catalog provides indicative information about possible risk alleles associated with the collected studies. Table 3 shows the data dictionary and sample entries of this cross-dataset, displaying data from the genetic literature along-side the GWAS data associated to the same rsID, such as Mapped Trait and Strongest Risk Allele. The resulting dataset comprises 385 unique genes with mapped data. Additionally, it includes 126 distinct MeSH terms, 316 mapped GWAS traits, and identifies 1,211 strongest SNP-risk alleles. This structured dataset aligns with our objective to facilitate precise and targeted retrieval in the nutrigenetics domain.

**Table 3:**
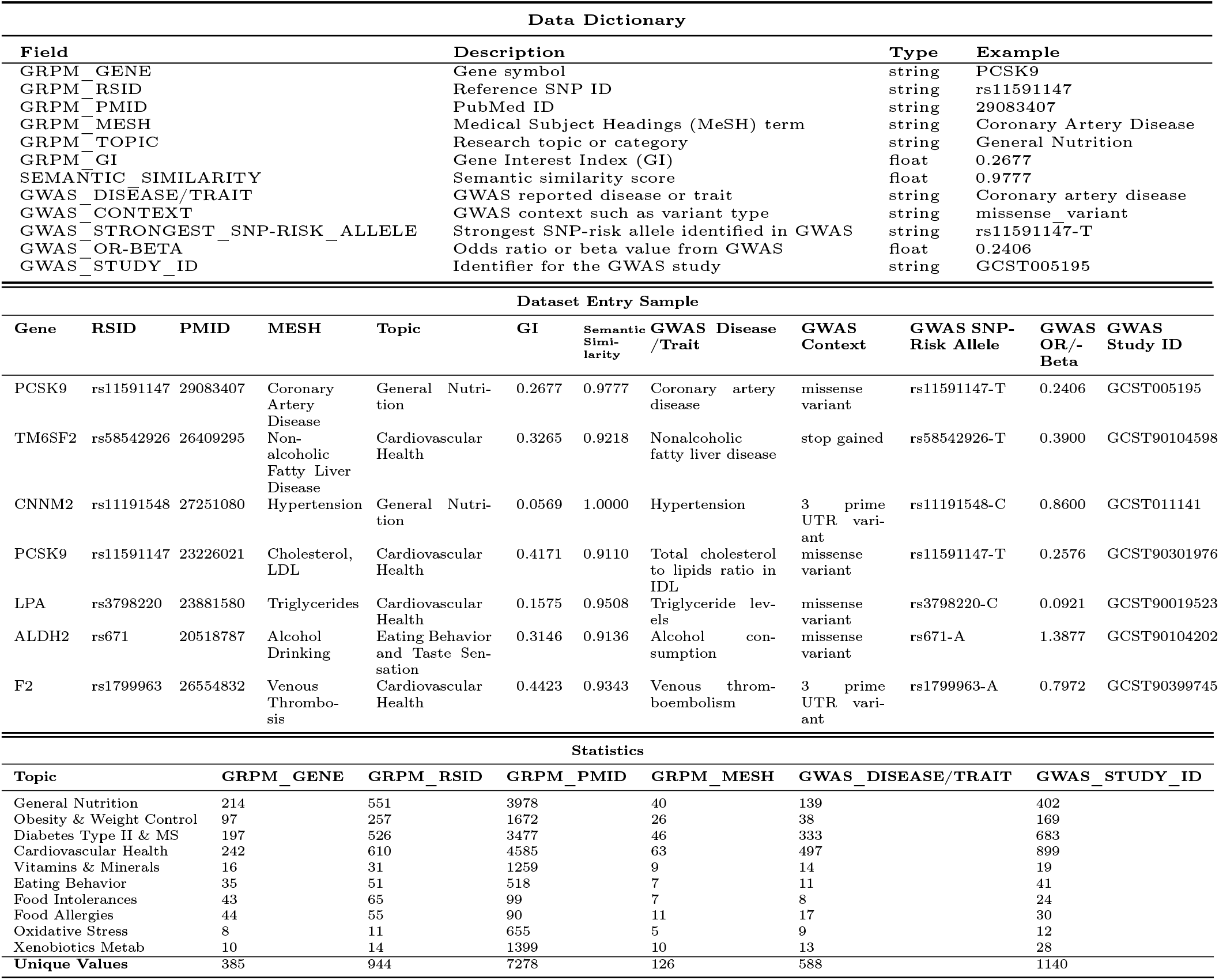
Nutrigenetic-GWAS Cross-Dataset description and statistics.

### 3.1 Data Analyisis

To elucidate the relationships within our dataset, we analyzed the prevalence of key genes and their associated MeSH terms and PMIDs across the ten nutrigenetic topics investigated in our research. This analysis, detailed in more detail in the Supplementary Material (see Figure S2), provides a comparative framework to assess the relevance of genes for various nutrition-related traits. In Figure S2, the richness of associated MeSH terms and PMIDs for the top 50 genes is highlighted, enabling an enhanced understanding of gene relevance within each topic.

To evaluate the extent of data overlap from the ten nutritional topics, we constructed co-occurrence matrices. Figure 4 illustrates the co-occurrence patterns among genes, rsID, PMID, and MeSH terms from the resulting nutrigenetic dataset. In this correlation analysis, we computed relative values for each topic by evaluating the ratio of shared entities to the total number of entities. This methodology allows for a detailed examination of co-occurrence within specific categories, facilitating a nuanced understanding of relationships and interactions. Remarkably, the correlation matrix in Figure 4 reveals substantial overlap between various MeSH term lists, such as the topic of ‘obesity’, which shares approximately 80% of its items with the topic of ‘diabetes’ and ‘cardiovascular health’, demonstrating a significant 40-60% overlap in this cluster of three categories.

**Figure 4:**
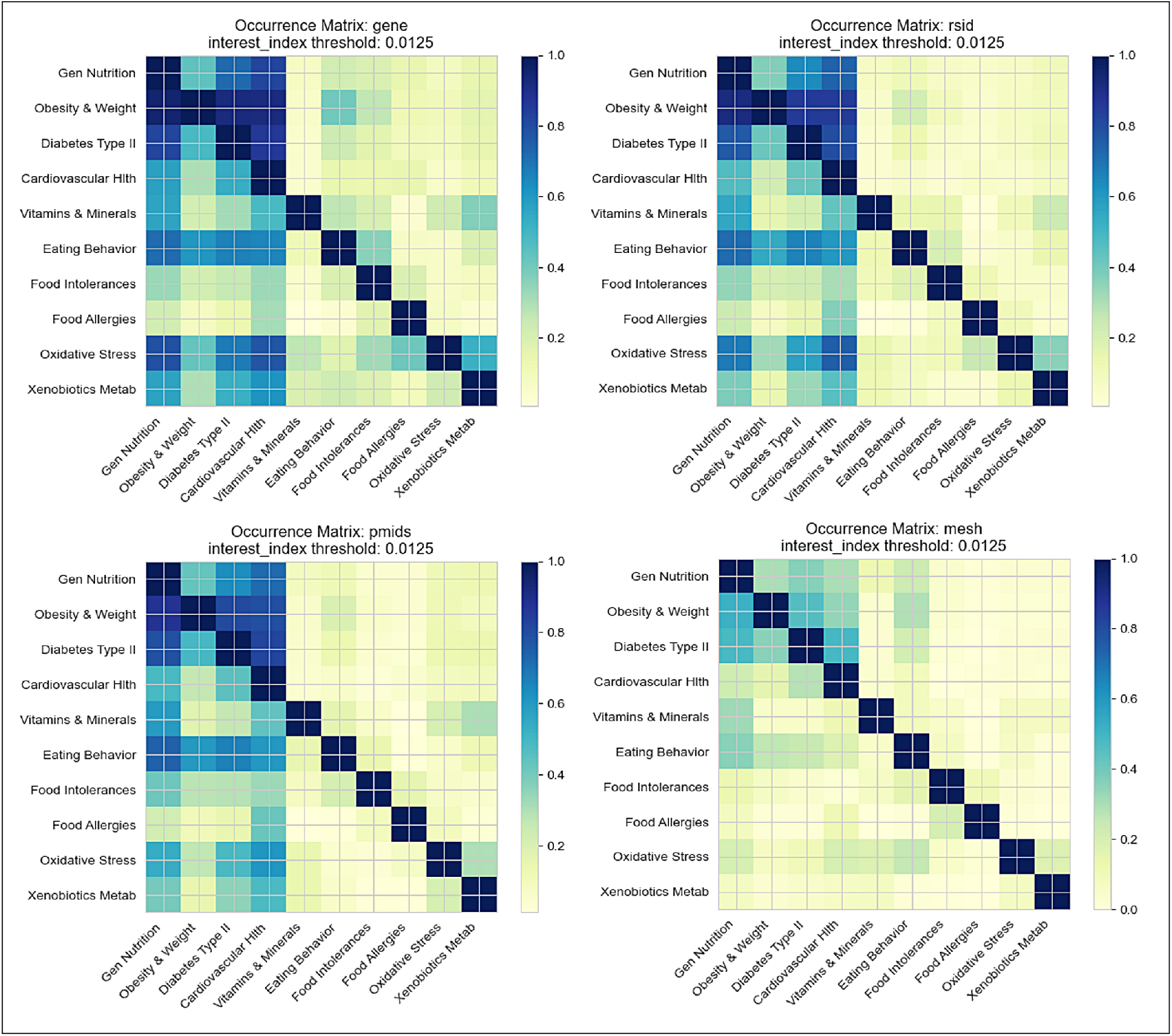
Co-occurrence patterns observed among genes, rsIDs, PMIDs, and MeSH terms in the refined dataset (GI > 0.0125) across ten nutritional topics. The color coding indicates the degree of overlap between datasets, normalized by the total size of each dataset.

### 3.2 Method Validation

Figure 5a shows the 50 most interesting genes for the topic *“Vitamin and Micronutrient Metabolism”* compared to the values of interest on results obtained from 5 other nutrigentic topics. The genes extracted using our method show specificity for the topic used as a reference. This behavior suggests that our method can identify genes related to that particular nutritional aspect. It can be seen in Figure 5a that some of the genes have higher GI in other topics than the one taken as a reference in the plot, meaning they are more closely associated with other nutritional features or specific biological processes. This observation suggests the complexity of gene regulation in nutrient metabolism and underscores the importance of considering a broader range of nutritional MeSH terms to gain a comprehensive understanding of the biological system under consideration. Supplementary Materials provide additional GI comparison results, showcasing the comparison among another MeSH list utilized in our study (Figures S3, S4). To ensure the accuracy and reliability of the data collected, we compared the results obtained with biologically consistent MeSH queries with those obtained with 20 random MeSH queries^6^ of the same size. Figure 5b provides an example of the comparison results in the *“General Nutrition”* topics. Another example is shown in Supplementary Materials (S4).

**Figure 5:**
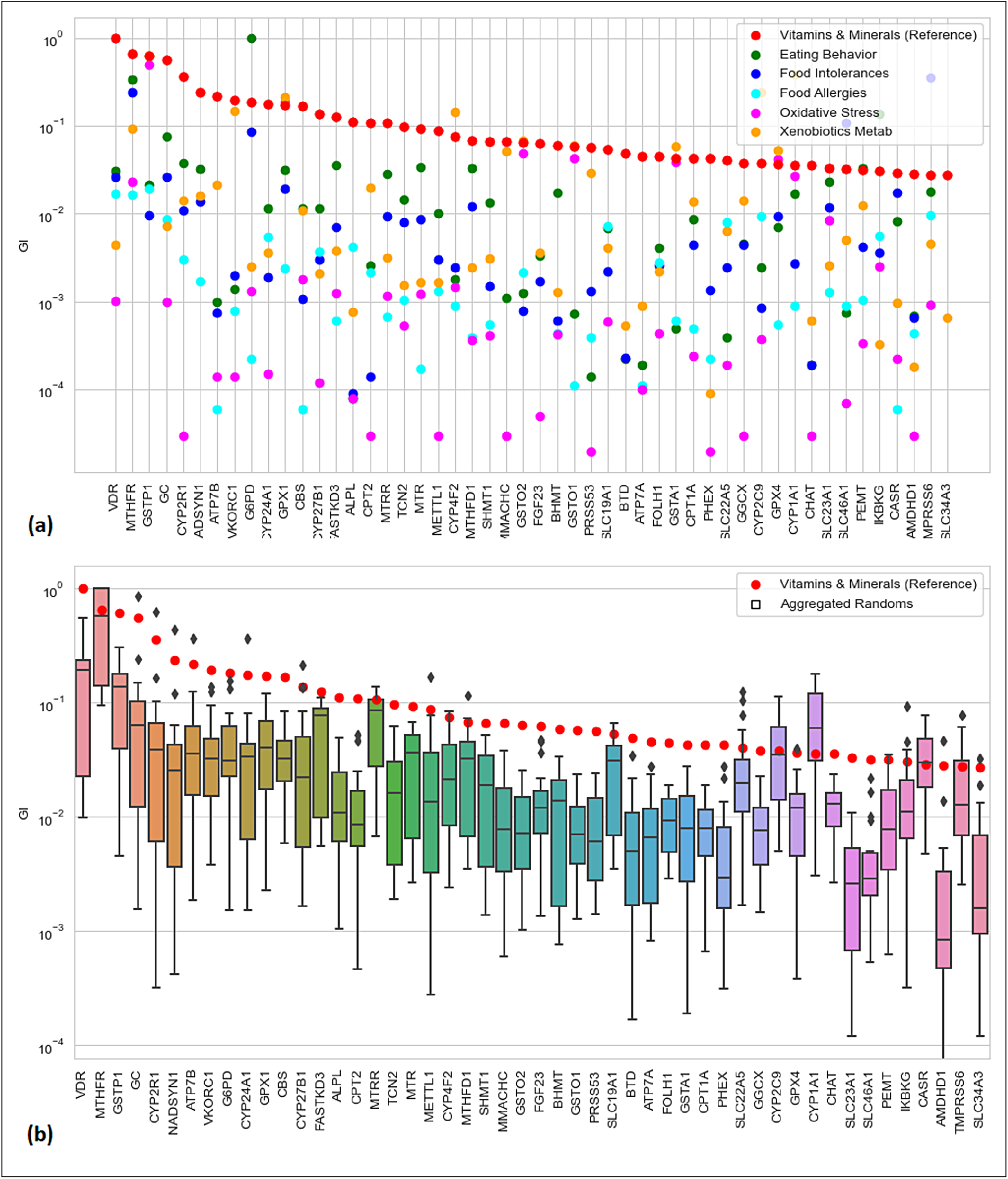
(a) Comparison of GI for the top 50 genes on the *“Vitamin and Micronutrient Metabolism”* topic with GI obtained from 5 different nutritional topics. (b) Comparison between GI obtained from the *“General Nutrition”* list and GI obtained using 20 randomly generated MeSH queries of the same size represented as boxplots. In both graphs the y-axis is logarithmic; genes are ordered by decreasing interest relative to the reference list.

## 4 Discussion

Understanding how genetic variations influence individual nutritional requirements, metabolism, and health outcomes is crucial for developing personalized nutrition interventions [18]. By considering an individual’s genetic profile, healthcare operators and nutritionists can provide tailored dietary advice, optimizing nutrient bioavailability, and promoting better health outcomes, thus preventing chronic diseases such as obesity [19], diabetes [20], or cardiovascular diseases [21].

Personalized healthcare and prevention strategies are central to advancements in translational bioinformatics [22]. The creation of sophisticated computational methodologies and tools for aggregating and analyzing data from diverse sources significantly enhances data integration, enabling the transformation of biomedical findings into tailored nutritional guidance and precision nutrition strategies for disease prevention [23, 24].

As remarked by Floris et al., the current approach adopted by many companies in nutrigenetic counseling still relies on a limited set of genes and polymorphisms for genetic testing and counseling [25]. However, as sequencing costs continue to decrease and sequencing technologies become more accessible than before, it is no longer justifiable to base nutrigenetic panels on a small number of genetic markers [25]. Using a limited set of genes and polymorphisms may overlook significant genetic variations that affect an individual’s response to nutrients and dietary patterns. Our study addresses the limitations of current approaches in nutrigenetics by consolidating and standardizing information on genetic polymorphisms associated with nutrition. Our nutrigenetic dataset offers a broader scope and coverage, improving the global understanding of the interplay between genetics and nutrition-related traits.

The resulting dataset is an integrated resource for diverse data sources related to genetic polymorphisms associated with nutrition. This resource enables efficient retrieval and analysis, facilitating comprehensive research in nutrigenomics. It is essential to highlight that this method’s potential applications extend beyond the scope of our study. Our approach can be employed to gather specific genetic polymorphisms associated with various health or biological dysfunctions, empowering healthcare practitioners to tailor interventions based on an individual’s genetic profile [26].

In our nutrigenetic dataset, the heterogeneity of MeSH term representation across nutrigenetic topics suggests that some topic encompass a wider array of aspects within nutrition, while others are more specialized with a focused scope. Notably, the genes highlighted in our study, as derived from MeSH queries (as detailed in Table 2), are prominent in nutrigenetics research. These genes, frequently cited in the literature, consistently appear across multiple MeSH queries, underscoring their significance regarding nutrition-related traits.

From a broader standpoint, the analysis revealed a high degree of overlap between genes associated with specific nutritional topics (Figure 4). For example, ‘obesity’ exhibit high overlap with the ‘diabetes’ and ‘cardiovascular health’, indicating shared genetic polymorphisms and pathways. This finding is not surprising given the close relationship between obesity, diabetes, and cardiovascular health, as these conditions often coexist and share common genetic and physiological factors. The high overlap suggests that shared genetic polymorphisms and pathways may be involved in these conditions. Conversely, the lower degree of overlap between specific topics could be attributed to their specificity, focusing on more specific biological processes or conditions with distinct genetic underpinnings. This behavior can be attributed to several factors. Firstly, these topics may be described by fewer MeSH terms, leading to a narrower focus and less overlap with others. Secondly, these topics may pertain to more specific biological processes or conditions with distinct genetic underpinnings than the broader conditions captured by the others. The integration of the nutrigenetic dataset with the GWAS data corroborated our findings by identifying potential risk alleles linked to specific genetic polymorphisms. Utilizing semantic similarity techniques, powered by the BioBERT language model, revealed a consistent alignment between MeSH terms associated with each PMID and the corresponding GWAS traits (Table 3). This congruence affirms that genetic associations identified through literature-based MeSH queries are reinforced by empirical evidence from GWAS studies.

To evaluate the validity and reliability of our results, we conducted a comparative analysis of the top-scoring genes across different nutritional queries, as depicted in Figure 5a. This validation step effectively demonstrated the capability of our method to accurately pinpoint genes pertinent to distinct nutritional topics. Additionally, we observed some genes exhibiting high relevance scores across multiple nutritional contexts, underscoring the intricate nature of gene regulation in nutrient metabolism and the necessity of considering various nutritional dimensions.

To validate the accuracy and reliability of our dataset, we assessed the outcomes generated from biologically coherent MeSH queries against those derived from random, non-cohesive queries (Figure 5b). Our analysis underscored that biologically consistent MeSH queries yielded significant and contextually relevant results for the nutritional aspects under investigation. In contrast, random MeSH queries produced non-significant outcomes with no meaningful associations. This comparison underscores the necessity of employing biologically pertinent MeSH queries for extracting and prioritizing data that is genuinely relevant to the specified biomedical domain.

However, it is essential to acknowledge the limitations of this approach. One limitation of our approach is the reliance on available literature and databases. The accuracy and reliability of the build resource depend on the quality and completeness of the data retrieved from various sources, as well as the accuracy of the data structuring and integration process. Our method relies on data from Medline studies, which may be subject to publication bias [27]. The data quality and consistency of retrieved data heavily depend on the quality of the original studies and the curation process. Despite efforts to ensure data quality, inconsistencies, errors, and biases in the original studies may still be present in the constructed dataset.

Moreover, our dataset is limited to the data available in the LitVar database, GWAS-Catalog, and the other sources used in our study [28]. As a result, it may not encompass all potential genetic polymorphisms associated with nutrition-related traits. Relying on available literature and data collection databases has limitations [29]. Despite our efforts to minimize MeSH attribution bias, the dataset could not contain all the relevant literature. Inconsistencies, errors, and biases in the original studies may be transferred to the constructed dataset. Finally, the dataset may cover only some populations and ethnicities, which could limit its applicability to diverse populations with different genetic backgrounds [30].

Furthermore, it is essential to acknowledge the complex and multifactorial nature of gene-environment interactions, including dietary factors [31]. While our dataset captures a subset of the possible interactions, it may not encompass their full complexity. In interpreting the associations between genetic polymorphisms and nutrition-related traits, it is crucial to consider other factors, such as environmental influences, epigenetic modifications, and gene-gene interactions [32]. Therefore, the complexity of gene-environment interactions, including interactions with dietary factors, requires further investigation beyond the scope of this research. Moreover, to improve the validity of the study results, it is essential to assess the quality and scientific validity of the literature sources through established criteria. Future research could follow the scientific validity assessment criteria described by Grimaldi et al.[33] to ensure the reliability of individual sources.

In considering the dataset’s future evolution, the framework is designed to accommodate ongoing advancements in nutrigenetics. The pipeline’s modular architecture allows for systematic updates with new data sources and literature, ensuring the dataset remains at the cutting edge of scientific discovery. However, future updates must be meticulously curated and validated to preserve the dataset’s integrity, acknowledging that emerging research may revise previously accepted insights. Implementing robust data governance structures is essential to mitigate inconsistencies and safeguard the dataset’s overall reliability.

In future work, this dataset can be harnessed for the application of advanced NLP techniques, such as more recent semantic analysis methodologies. Currently, this dataset has already been emloyed for data-driven topic modeling and graph-based semantic analysis, which helps to uncover underlying themes within the nutrigenetics literature [34]. By employing text mining techniques, particularly those utilizing pretrained language models, the analysis of these datasets can be significantly enhanced. This will facilitate the identification of complex interactions between genes and dietary factors, allowing for the detection of patterns and correlations as well as the development of predictive models [35].

## 5 Conclusion

Our study presents a comprehensive nutrigenetic dataset, constructed by integrating data from multiple sources using the MeSH ontology. This dataset is a valuable resource for exploring genetic polymorphisms associated with nutrition-related traits. By consolidating and standardizing genetic polymorphism data, our work aims to advance personalized nutrition interventions and contribute to the field of nutrigenomics.

The dataset, openly available, fills a significant gap in the existing resources in the field, providing a reliable and unified resource for investigating gene-diet interactions. It underscores the importance of standardized curation processes and highlights the role of translational bioinformatics in merging and analyzing information from diverse sources. By doing so, it facilitates comprehensive research in nutrition and genetics, offering a practical tool for researchers and nutritionists alike. We hope this dataset will serve as a foundational resource for future nutrigenetic studies and help in the development of personalized nutrition strategies based on genetic insights.

## Data Availability

Data and code are available on Zenodo and GitHub, respectively. URLs are provided in the "Data Availability Links section.

https://zenodo.org/record/8205724

https://github.com/johndef64/GRPM_system

## Author contributions

GMDF: Writing - original draft, Conceptualization, Data curation, Software, Visualization, Writing – review & editing; MM: Writing - original draft, supervision, Investigation, Writing – review & editing; AP: Supervision, Writing – review & editing, Validation; TA: Supervision, Writing – review & editing, Validation; BHM: Writing - original draft, Conceptualization, Formal analysis, Validation, Writing – review & editing; VC: Funding acquisition, Conceptualization, Supervision, Writing – review & editing.

## Acknowledgements

Our research was supported by Federazione Nazionale Degli Ordini dei Biologi.

## Footnotes

1 https://github.com/johndef64/GRPM_system

2 https://www.ncbi.nlm.nih.gov/research/litvar2/api

3 https://pypi.org/project/nbib/

4 The complete MeSH dataset can be downloaded at https://www.nlm.nih.gov/mesh/meshhome.html

5 https://www.ebi.ac.uk/gwas/docs/file-downloads

6 Containing 450 random terms each, based on 21,705 MeSH in the GRPM dataset.

