## Supplemental materials for "Computational Strategies in Nutrigenetics: Constructing a Reference Dataset of Nutrition-Associated Genetic Polymorphisms"

### 1 Supplementary Tables

Table S1: Nutrition-related Topics

| **Topic** | **Description** | **MeSH count** |
|---|---|---|
| *General Nutrition* | A broad range of topics related to nutrition, including dietary patterns, nutrient requirements, nutritional status, and the impact of nutrition on overall health and well-being. | 413 |
| *Obesity, Weight Control, and Compulsive Eating* | Terms related to weight management, including obesity, weight loss strategies, and disorders such as binge eating or compulsive overeating. | 243 |
| *Cardiovascular Health and Lipid Metabolism* | Terms related to nutrition and cardiovascular health, including the impact of dietary factors on lipid metabolism, cholesterol levels, and the prevention of cardiovascular diseases. | 319 |
| *Diabetes Mellitus Type II and Metabolic Syndrome* | Terms related to type II diabetes and metabolic syndrome. Including dietary interventions, glucose metabolism, insulin resistance, and related complications. | 528 |
| *Vitamin and Micronutrients Metabolism and Deficiency-Related Diseases* | Terms related to the metabolism of essential vitamins and micronutrients, the impact of deficiencies on health and the development of associated diseases. | 175 |
| *Eating Behavior and Taste Sensation* | Terms related to individual eating behaviors, including factors influencing food choices, taste preferences, satiety, and appetite regulation. | 292 |
| *Food Intolerances* | Terms related to adverse reactions to specific foods, such as lactose intolerance or gluten sensitivity. Explores the genetic and physiological factors underlying food intolerances and their impact on dietary choices. | 145 |
| *Food Allergies* | Examines the genetic basis of food allergies, the identification of allergenic components, and strategies to manage allergic reactions through diet. | 65 |
| *Diet-induced Oxidative Stress* | Explores the relationship between dietary factors and oxidative stress and investigates the impact of diet on oxidative stress levels and its health implications. | 77 |
| *Xenobiotics Metabolism* | Focuses on the metabolism of foreign substances (xenobiotics) in the body, including drugs, environmental toxins, and dietary components. | 170 |

### 2 Supplementary Figures

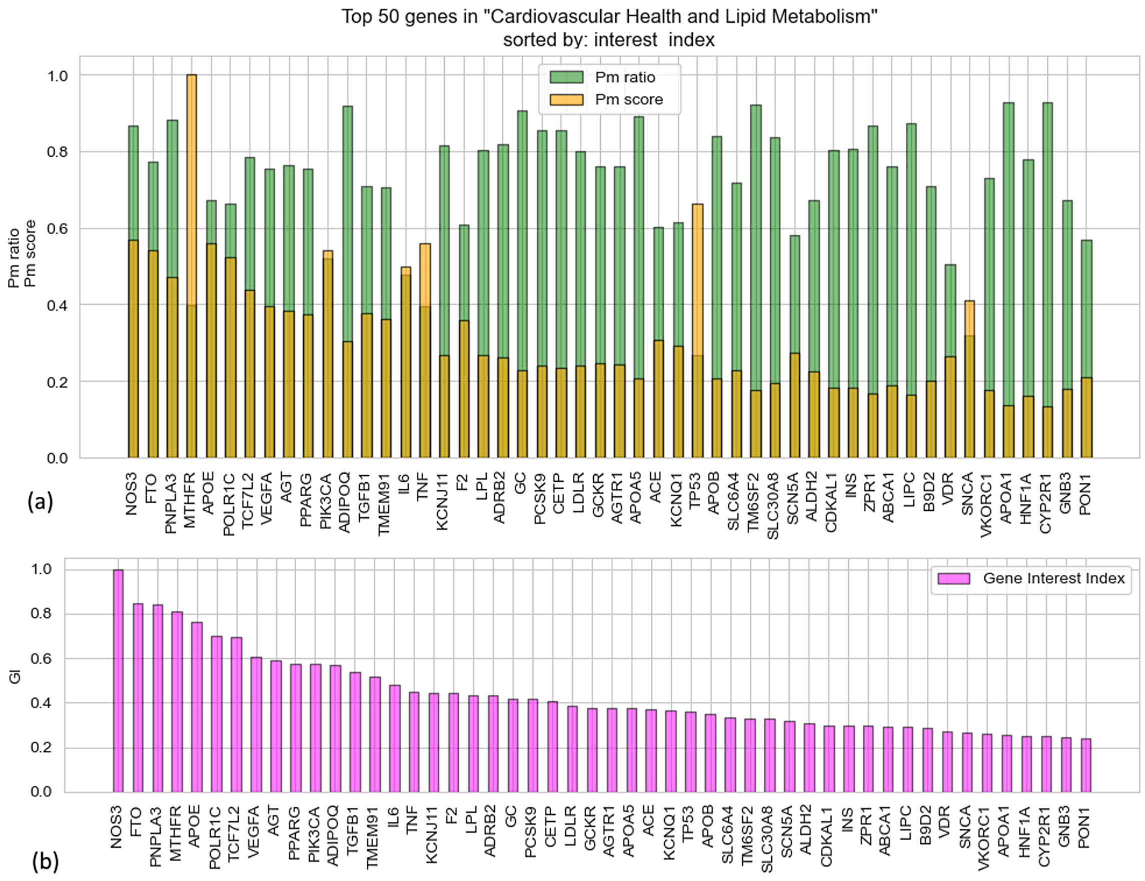


Figure S1: Another example of gene prioritization through GI taking as reference the results from the "Cardiovascular health" MeSH query. Panel (a) shows the matching PMID ratio and overall matching PMID score. Panel (b) displays the gene relevance sort achieved with the GI.

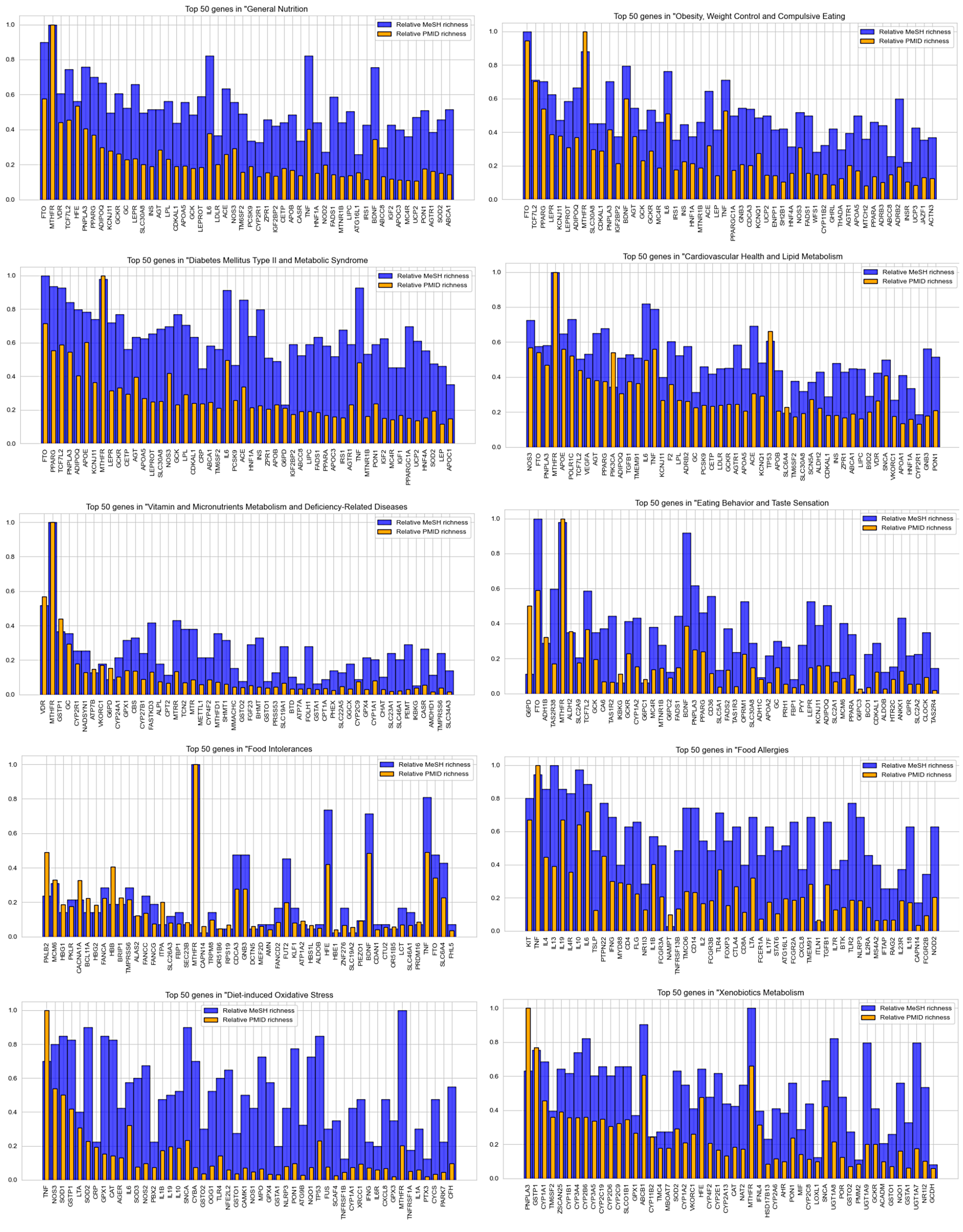


Figure S2: Overview of the fifty most interesting genes with their relative MeSH and PMID values on the ten nutrient lists used in the study. The MeSH column (blue) displays the relative richness of the associated MeSH terms used, while the PMID column (orange) shows the relative richness of papers associated with each gene.

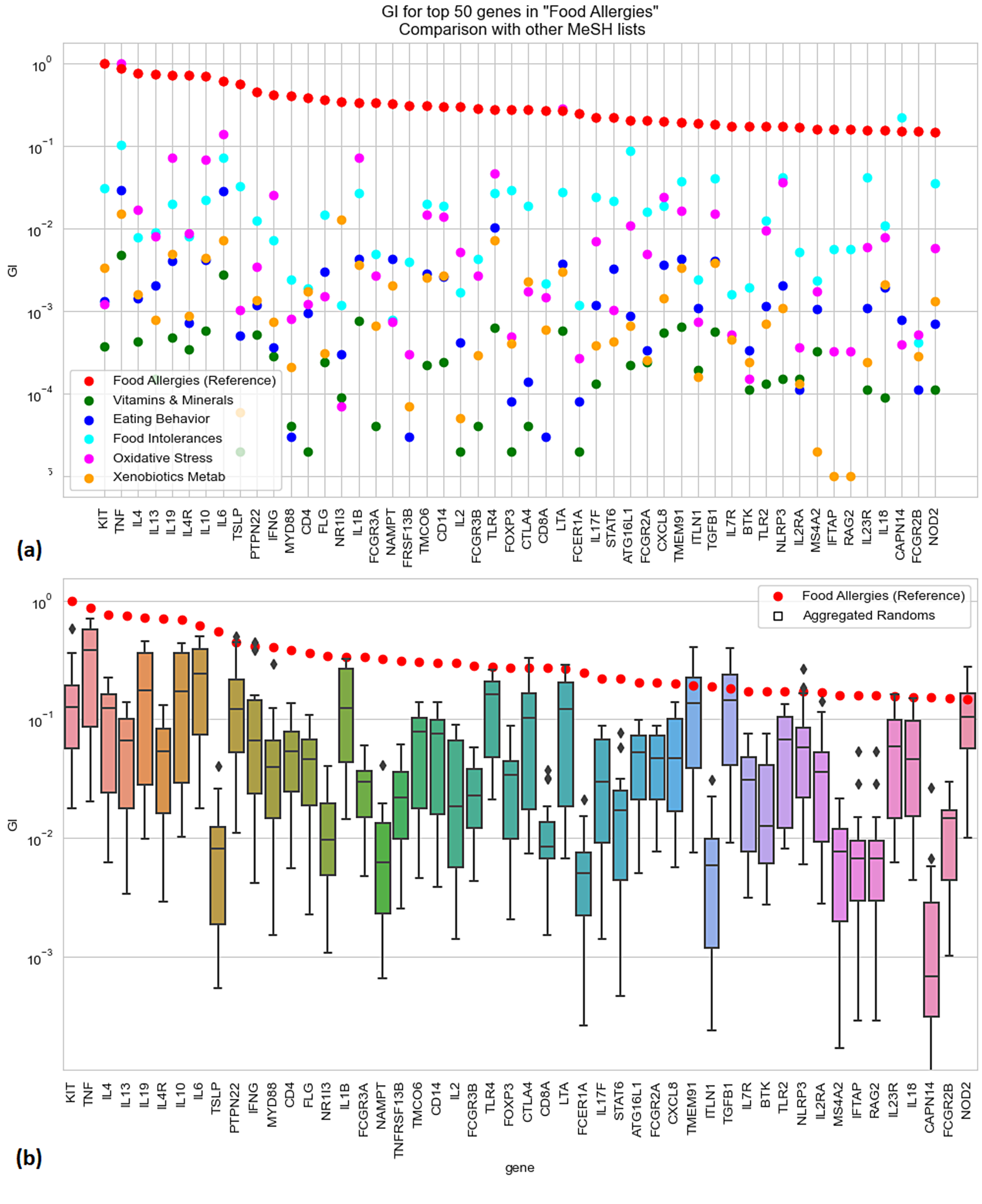


Figure S3: Gene Interest Index for the top 50 genes on the "Xenobiotics Metabolism" MeSH list. (a) Comparison with GI obtained on other five different lists of nutrition MeSH terms. The y-axis is logarithmic; genes are ordered by decreasing interest relative to the reference list of this plot.

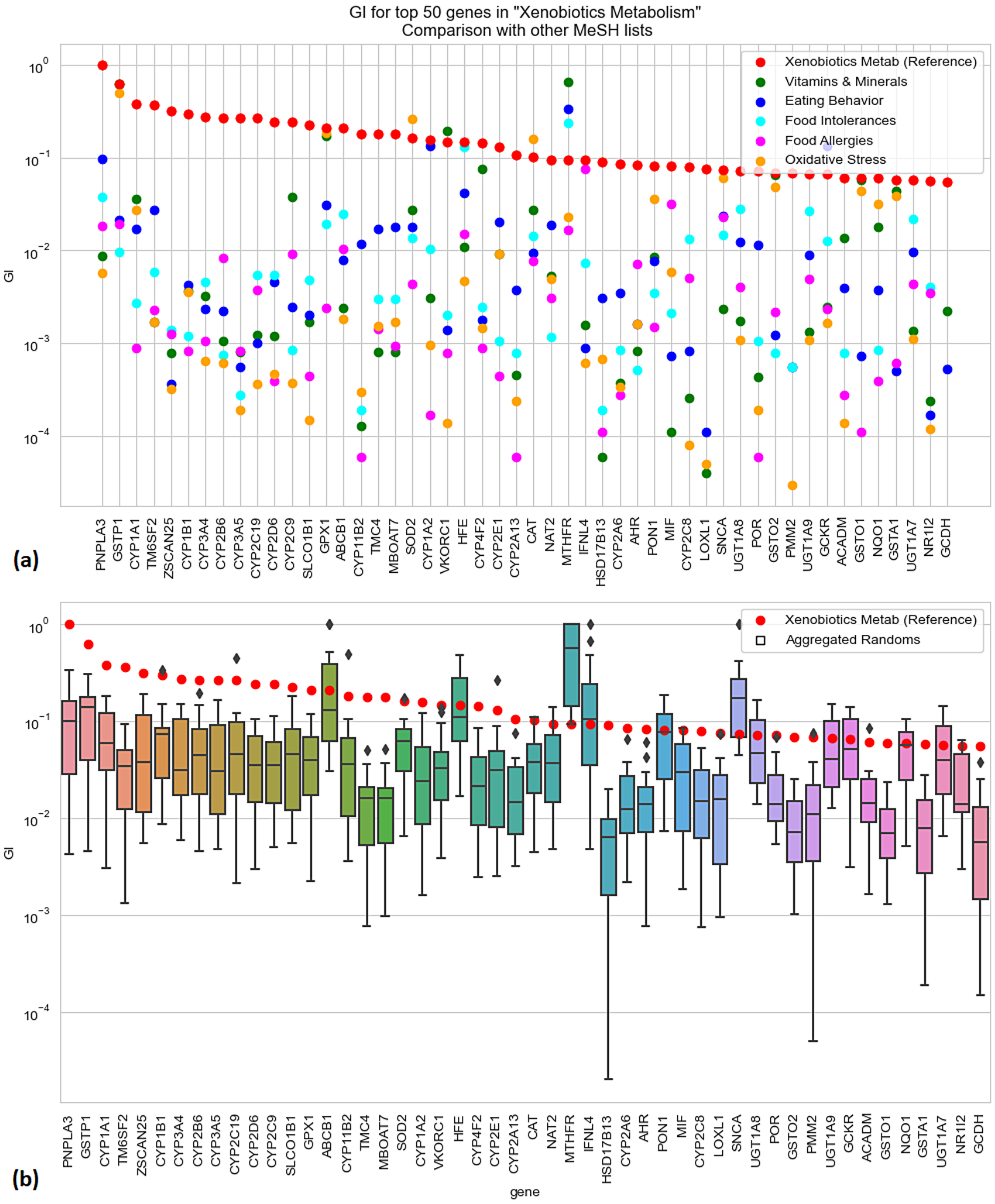


Figure S4: Gene Interest Index for the top 50 genes on the "Cardiovascular Health" MeSH list. Comparison of GI obtained using 20 randomly generated MeSH lists represented as boxplots. The y-axis is logarithmic; genes are ordered by decreasing interest relative to the reference list of this plot.
